# Segregation of dynamic resting-state reward, default mode and attentional networks after remitted patients transition into a recurrent depressive episode

**DOI:** 10.1101/2022.09.02.22279550

**Authors:** Sonsoles Alonso Martinez, Anna Tyborowska, Nessa Ikani, Roel J. Mocking, Caroline A. Figueroa, Aart H. Schene, Gustavo Deco, Morten L. Kringelbach, Joana Cabral, Henricus G. Ruhé

## Abstract

**Introduction:** Recurrence in major depression disorder (MDD) is common, but neurobiological models capturing vulnerability for recurrences are scarce. Disturbances in multiple resting-state networks have been linked to MDD, but most approaches focus on stable (vs. dynamic) network characteristics. We investigated how the brain’s dynamical repertoire changes after patients transition from remission to recurrence of a new depressive episode.

**Methods:** Sixty drug-free, MDD-patients with ≥2 episodes underwent a baseline resting-state fMRI scan when in remission. Over 30-months follow-up, 11 patients with a recurrence and 17 matched-remitted MDD-patients without a recurrence underwent a second fMRI scan. Recurrent patterns of functional connectivity were characterized by applying leading eigenvector dynamics analysis (LEiDA). Differences between baseline and follow-up were identified for the 11 non-remitted patients, while data from the 17 matched-remitted patients was used as a validation dataset.

**Results:** After the transition into a depressive state, the reward and a visuo-attentional networks were detected significantly more often, whereas default mode network activity was found to have a longer duration. Additionally, the fMRI signal in the areas underlying the reward network were significantly less synchronized with the rest of the brain after recurrence (compared to a state of remission). These changes were not observed in the matched-remitted patients who were scanned twice while in remission.

**Conclusion:** These findings characterize the changes that are specifically associated with the transition from remission to recurrence and provide first evidence of increased segregation in the brain’s dynamical repertoire when a recurrent depressive episode occurs.

## Introduction

Major Depressive Disorder (MDD) is a severe and highly prevalent disease affecting 5% of the global population (Institute of Health Metrics and Evaluation, 2019). An important factor in the high personal and societal impact of MDD is the high risk of recurrence (Bockting, Spinhoven, Wouters, Koeter, and Schene, 2009; Hardeveld, Spijker, De Graaf, Nolen, and Beekman, 2010). Nevertheless, neurobiological investigations of the recurrence of depressive episodes remain scare. This study addresses the critical issue of how network dynamics change when patients transition from a remitted to a recurrent depressed state.

MDD has been associated with disturbances in multiple resting-state networks, including the default mode network (DMN), salience and frontal networks —regulating cognitive control and attention (Kaiser, Andrews-Hanna, Wager, and Pizzagalli, 2015; Menon, 2011; Mulders, van Eijndhoven, Schene, Beckmann, and Tendolkar, 2015). Recent studies have further shown that MDD may be related to alterations in large-scale functional connectivity (FC) between these networks (Li et al., 2020; Liu et al., 2020). Specifically in remitted depressed patients, functional hyper-connectivity in e.g. DMN and dorsal attention network, as well as within and between salience and executive control networks have been reported (Liu et al., 2021).

The majority of research conducted in MDD has largely focused on discerning differences between patients and healthy controls or predicting vulnerability for relapse from baseline assessments. In this respect, dominance of activity within the DMN has been associated with MDD recurrence and related to rumination (Lythe et al., 2015; Marchetti, Koster, Sonuga-Barke, and De Raedt, 2012), whilst decreased within network FC of the DMN has been shown to predict more depressive symptoms within a 2 year follow-up period (Blank et al., 2021). Another study points to the neuro-progressive nature of MDD, with remitted depressed patients (compared to first episode patients) showing hyper-connectivity in a wide range of networks, including the DMN (Liu et al., 2021). However, a within subject investigation of changes in large-scale FC during the transition into a depressive episode, to our knowledge, has never been investigated.

A recent meta-analysis suggests limited convergence of findings across resting-state measures (Gray, Müller, Eickhoff, and Fox, 2020), despite previous reports to the contrary (Kaiser et al., 2015). Heterogeneity in findings may be related to the fact that studies thus far have primarily applied static resting state measures, which do not provide insights on the progression of brain network connectivity over time. It is, therefore, crucial to use a dynamic approach that assesses the integration and segregation (Deco, Tononi, Boly, and Kringelbach, 2015) of time-varying neurocognitive brain networks (Barrett and Satpute, 2013; Bressler and Menon, 2010; Deco and Kringelbach, 2014; Yarkoni, Poldrack, Nichols, Van Essen, and Wager, 2011) in order to adequately capture the neural correlates of MDD and its recurrence.

A promising avenue in this regard is dynamic FC, based on growing evidence that neural activity at rest is not stable, but slowly fluctuates through varying, but repeating states (Cabral, Kringelbach, and Deco, 2017). For example, a recent study in MDD-patients has shown disrupted organization of dynamic brain networks with respect to higher variability and lower consistency in FC between time-points, as compared to healthy controls (Long et al., 2020).

A novel method for investigating dynamic FC is Leading Eigenvector Dynamics Analysis (LEiDA), which identifies whole-brain phase-locking patterns in fMRI signals at every time-point, that are clustered into repeating FC states (Cabral et al., 2017). Each FC state reveals synchronization within a specific set of brain areas and is characterized by its probability of occurrence (fractional occupancy) and duration (lifetime). In comparison to other analytical tools, LEiDA extends from measures of connectivity or correlation by considering also the phase-shifts between brain regions and describes discrete instead of overlapping states in time (Kringelbach and Deco, 2020). Dynamic characteristics of FC states derived from LEiDA have been related to cognitive performance (Cabral et al., 2017), emotionality (Stark et al., 2020), depressive symptoms (Alonso Martínez, Deco, Ter Horst, and Cabral, 2020), trait self-reflectiveness (Larabi et al., 2020), body dysmorphic disorder (Wong et al., 2021) and distinct mood states in remitted MDD (rMDD) patients (Figueroa et al., 2019), which reinforces its sensitivity to both clinical and pre-clinical psychiatric symptoms.

The goal of the current study was to investigate how dynamic FC states change when patients shift from remission to recurrence of a new depressive episode. We hypothesized that patients experiencing a recurring depressive episode would show changes in fractional occupancy and lifetime of FC states of DMN, attentional and salience networks, compared to a remitted state.

## Materials and Methods

### Participants

As part of a larger project investigating the neurobiology of recurrence of depressive episodes (Mocking et al., 2016); 62 drug-free (>4 weeks), rMDD-patients (age 35–65 years) with ≥2 episodes (according to the Structured Clinical Interview for DSM-IV Disorders (SCID)); underwent a baseline resting-state fMRI scan when in remission (Hamilton Depression Rating Scale (HDRS-17) score ≤7 for ≥8 weeks). Exclusion criteria were alcohol/drug dependency; psychotic or bipolar disorder; predominant anxiety disorder or severe personality disorder; electroconvulsive therapy within 2 months before scanning and current severe physical illness. MRI exclusion criteria were: incompatible implants or tattoos, claustrophobia, history of seizure or head injury, and neurological disorder. Participants were recruited from primary care, secondary mental health-care institutes, from previous studies and through advertisements in online and house-to-house newspapers and posters in public places.

Over a 30-month follow-up period, a second fMRI scan was obtained from patients reporting a recurrence. Overall, 55% of patients had a recurrence. Of these, in eleven patients we were able to capture the recurrent depressive episode in time and motivate the patient for a second scan during the depressive episode. Seventeen rMDD-patients without a recurrence; matched for age, sex, IQ and length of follow-up were also scanned a second time (Table 1). For more information on the full sample and recurrence rates see (Figueroa et al., 2019; Ruhe et al., 2019). Informed consent was obtained prior to participation; the study was approved by the local Medical Ethical Committee of AmsterdamUMC.

**Table 1.** Sample characteristics

|  |  | **Nonrecurrent** | **Recurrent** |  |
| --- | --- | --- | --- | --- |
|  |  | **(n=17)** | **(n=11)** |  |
| Sex | Women | 13 | 9 | $\chi^2(1)=.133$ |
|  | Men | 4 | 2 |  |
| Age | | 53.5 (6.9) | 50.5 (5.75) | $t(26)=1.23$ |
| IQ | | 107 (9.21)<sup>#</sup> | 108 (8.04)<sup>^</sup> | $t(23)=.084$ |
| Episodes in the past (number) | | 6.76 (11.8) | 11.40 (18)<sup>^</sup> | $U=108$ |
| Months since Baseline | | 11.6 (5.86) | 10.2 (6.27) | $t(26)=.601$ |
| Completed FU time-point at 2<sup>nd</sup> MRI | | 2.18 (1.38) | 2.45 (1.57) | $U=106$ |
| HDRS Baseline | | 3.69 (2.85) | 6.11 (5.49) | $U=117$ |
| HDRS at 2<sup>nd</sup> MRI | | 3.73 (3.64) | 20.4 (5.38) | $U=185$ ** |

Values indicate the mean and standard deviation.
FU, follow-up; HDRS, Hamilton Depression Rating Scale
\*\* $p < .001$
<sup>#</sup>n=15; <sup>^</sup>n=10

### Image acquisition

A 3 Tesla Philips Achieva XT scanner (Philips Medical Systems, Best, the Netherlands), with a 32-channel SENSE head coil, was used to obtain the images. A high-resolution T1-weighted 3D structural image was acquired using fast-field echo (FFE) for anatomical reference (220 slices; TR: 8.3 ms; TE: 3.8 ms; FOV: 240×188; 240×240 matrix; voxel size: 1×1×1 mm). Functional images were acquired with a T2*-weighted gradient echo-planar imaging (EPI) sequence. Participants were instructed to close their eyes and stay awake. The scan comprised 208 volumes of 37 axial-slices (TR: 2000 ms; TE: 27.6 ms; FOV: 240×240; 80×80 matrix; voxel size: 3×3×3 mm), oriented parallel to the AC–PC transverse plane and acquired in ascending order with a gap of 0.3 mm.

### Image preprocessing and analysis

#### Image preprocessing

fMRI data was preprocessed using FSL version 6.0 (FMRIB, Oxford, UK). To control for T2* equilibration effects, the first four volumes were discarded. Images were aligned to the first scan using rigid body transformations, spatially smoothed with a 5mm FWHM Gaussian kernel, grand-mean intensity normalization by a single multiplicative factor, and denoised using a non-aggressive ICA-AROMA procedure (Pruim et al., 2015). Model nuisance effects were regressed out from the preprocessed images with a linear model. These included 24 head-motion parameters: 6 motion regressors, their derivatives, and squared terms of each (Caballero-Gaudes and Reynolds, 2017; Friston, Williams, Howard, Frackowiak, and Turner, 1996). Brain extraction of T1 images was performed using the FSL anat preprocessing tool, resulting in bias-corrected images, subsequently segmented into white matter and CSF. Subject-specific white matter and CSF masks were thresholded at 95% probability and coregistered to the functional images. Mean signal intensities were extracted per volume and regressed from the preprocessed fMRI images along with the motion regressors. High pass filtering with a cut-off of 100s was performed on the residual images from the linear model. Images were normalized to the Montreal Neurological Institute template (MNI152) using linear (FLIRT; Jenkinson et al., 2002; Jenkinson and Smith, 2001) and nonlinear (FNIRT; Andersson et al., 2007) transformations via boundary-based registration (BBR; Greve & Fischl, 2009).

Time series were extracted using an 80-region Mindboggle parcellation, given its clinical relevance. The Desikan-Killiany-Tourville (DKT) labeling protocol (Desikan et al., 2006) was used to parcellate each cortical hemisphere into 31 anatomical regions and 9 subcortical regions. The FSL function, fslmeants, was used to calculate an average over voxels within each ROI to get the representative time courses. Lastly, temporal band-pass filtering was applied to detrend the time course signal and to retain frequencies between 0.01–0.1 Hz.

#### Leading Eigenvector Dynamics Analysis

We applied LEiDA to characterize recurrent phase-locking (PL) patterns in fMRI signals. This data-driven approach relies on the leading eigenvector of the phase coherence matrix at each single TR (Cabral et al., 2017). First, participant-specific sets of 80 ROI time courses were demeaned and Hilbert transformed to estimate the phase of the ROI signals (**Figure 1A**). The Hilbert transform expresses any given signal *x* in polar coordinates, i.e., *x = A* cos *θ*, where *A* is the instantaneous amplitude, and *θ* the instantaneous phase at a given time point. Then, at each time point, the phase coherence between 2 regions, *n* and *p*, can be calculated as the cosine of the phase differences as in the following equation:

**Figure 1.**
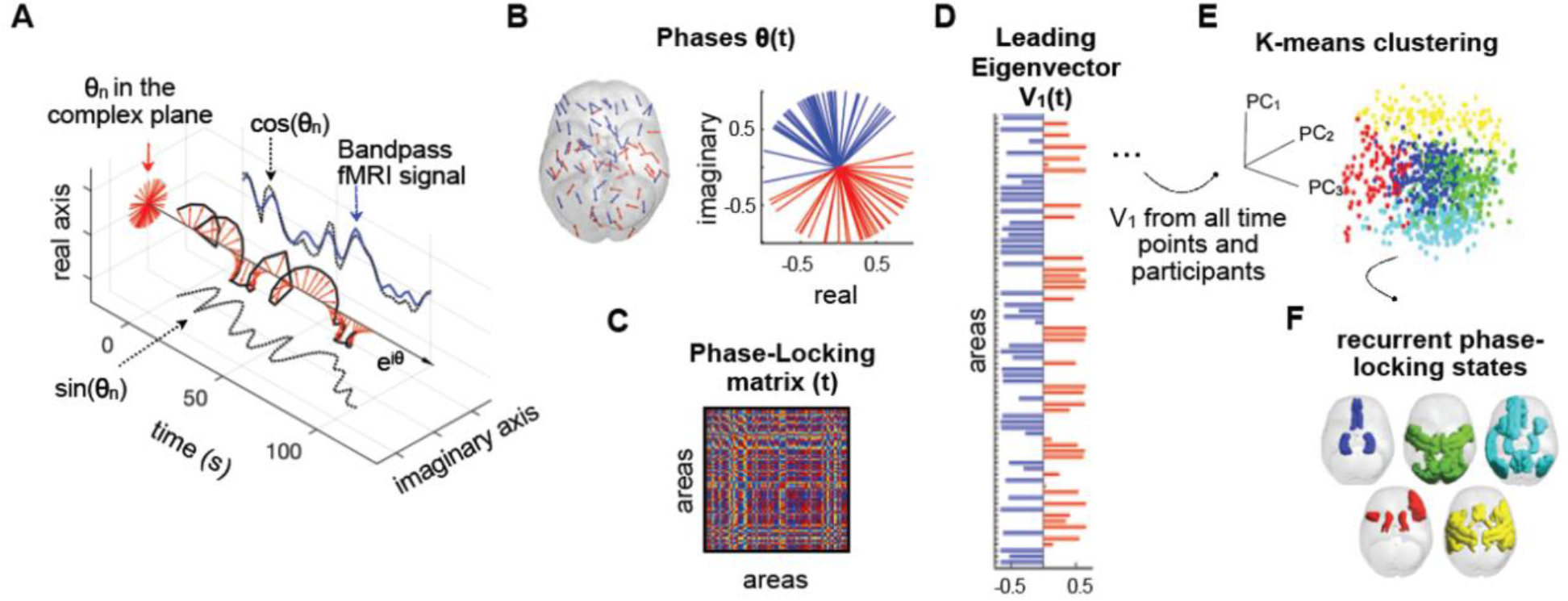
Detection of recurrent phase-locking (PL) patterns in fMRI signals. **A**) The fMRI signal is band-pass filtered between 0.01–0.1 Hz (blue) and Hilbert transformed into an analytic signal, whose phase can be represented over time (eiθ black arrow) and at each TR (red arrows). **B)** The phases in all N = 80 regions at a single TR are represented in the complex plane (right) and cortical space (left; arrows are placed at the center of gravity of each region n; the direction and color of the arrows indicate the sign of the corresponding element in the leading eigenvector V1(n,t) (red for positive and blue for negative). **C)** The PL matrix captures the phase alignment between each pair of regions. **D)** The leading eigenvector of the PL matrix at time t, V1(t) captures the main orientation of all phases. **E)** The leading eigenvectors obtained for each time point are concatenated over scans and subjects, and fed into a k-means clustering algorithm which divides the pool of data points into a predefined number of clusters k. **F)** Cortical representation of the PL-states (clusters). PL = phase-locking (right).

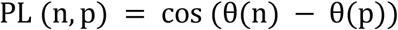

where PL takes values from 1 to −1, for signals changing in the same or opposite direction, respectively (**Figure 1B**). This process results in a time-resolved dynamic phase-locking (dPL) matrix with size *N*×*N*×*T*, where *N* (=80) is the number of brain regions and *T* (=204) is the number of recording frames in each scan (**Figure 1C**). The first and last volumes of each scan were removed to account for boundary distortions associated with the Hilbert transform. Next, the Leading eigenvector, *V1*, of each PL matrix was calculated (**Figure 1D**). The leading eigenvector captures the dominant instantaneous connectivity pattern, which substantially reduces data dimensionality from *N*×*N* to *N*×1.

#### Detection and characterization of recurrent PL-states

We employed a data mining approach that successively partitions the data searching for the partitions that are able to detect changes associated with the recurrence of a new MDD episode. Although an open question in dynamic FC research is the number of recurrent brain states, we here do not aim to identify the optimal number of PL-states explored during resting-state activity. Instead, we aimed to identify and characterize the dynamics of recurrent FC patterns that could potentially explain changes associated with the experience of a new depressive episode. As brain signatures related to MDD recurrence may arise at different levels of granularity, we used a *k*-means clustering algorithm which was run for 19 partition models by varying the number of clusters *k* from 2 to 20, with higher *k* resulting in more fine-grained configurations. For each *k*, the 4,488 leading eigenvectors (resulting from 11 subjects, 2 scans, 204 volumes each) were partitioned into *k* clusters (**Figure 1E**), resulting in *k* cluster centroids of *N*×1 dimensions, each representing a recurrent PL-state (**Figure 1F**). With the PL-states derived from the data of the 11 recurring patients, we applied these to the 17 matched nonrecurring, remitted patients used for the validation step. In these 17 nonrecurring rMDD-patients, 6,936 leading eigenvectors were obtained (17 subjects, 2 scans, 204 volumes); a *k*-means clustering with a single iteration, using the cluster centroids from the recurring group as ‘start vectors’ defining identical PL-states.

PL-states were characterized according to their fractional occupancy and lifetime. Fractional occupancy is calculated as the temporal proportion of epochs assigned to a given cluster centroid, that is, the proportion of time in which a PL-state was active. The lifetime of each PL-state is the mean number of consecutive epochs in the same state.

#### Phase-shifted signals between brain regions

Previous studies show that LEiDA captures relevant phase-shifts between brain regions (e.g., Cabral et al., 2017; Figueroa et al., 2019), which is often missed with typical measures of correlation. To further investigate the relevance of these phase-shifts, we obtained a continuous metric to estimate the phase shift of a given functional subsystem from the rest of the brain. Since each PL-state is represented by a Nx1 vector with positive and negative elements, we calculated the mean phase across regions with a positive sign and the mean phase across regions with a negative sign, and then calculated the angular difference between these 2 phase signals over time - obtaining a temporal signature capturing the degree of shift between the functional network detected in each PL-state and the rest of the brain. We averaged the angular difference over time for a metric of total phase shifting of a given functional network that can be compared between scan-sessions or subjects.

### Statistical analysis

Given the moderate number of patients involved, a repeated measures ANOVA investigating the group-by-time interaction was not justified. Therefore, differences in fractional occupancy and lifetime were statistically assessed between scans in the recurring group separately, using a nonparametric permutation-based paired t-test (5,000 permutations). In line with Figueroa et al. (2019), p-values were adjusted for each of the 19 partition models obtained by *k*-means clustering by controlling the false discovery rate (FDR) as proposed by Benjamini & Hochberg (1995).

As indicated above, with the PL-states identified in recurring patients, we repeated the analyses in the matched nonrecurring rMDD-patients, as a validation of the specificity of findings. Moreover, in a sensitivity analysis, we determined the reoccurrence of PL-states in the nonrecurring rMDD-patients separately. Therefore, the k-means clustering was applied directly on the 6,936 leading eigenvectors from the recurrent patients (from 17-subjects, 2-scans, 204-volumes), and the two sessions were compared using a nonparametric permutation-based paired t-test (5,000 permutations) for fractional occupancy and lifetime.

In the identified PL-states, post-hoc, we compared the decoupling metric between the two sessions. Again, we used permutation paired t-tests in the recurring and nonrecurring group separately.

## Results

Recurring and nonrecurring patients did not differ with respect to number of men/women, age, IQ, number of depressive episodes, months since baseline assessment, and follow-up time-point of the 2^nd^ MRI session. The groups were matched on HDRS scores at baseline, but as expected, differed on their HDRS scores during the follow-up scan, with recurrent patients having a significantly higher symptom score. See Table 1 for sample characteristics and statistics.

### Recurrence-related changes in PL-state fractional occupancy

Significant differences in fractional occupancy between baseline and follow-up occur in states characterized by fine-grained network configurations (**Figure 2**). Partition k=18 was the first partition model to return significant changes associated with the recurrence of a new MDD episode. Specifically, the average fractional occupancy of two states increased from remission to recurrence: PL-state k18c10 (*t*(10)=2.91; *p*-FDR=0.039, *d*=0.88), which comprises areas of the reward system, namely, basal ganglia and anterior cingulate cortex (ACC), and k18c15, (*t*(10)=3.14; *p*=0.004; *p*-FDR=0.039, *d* =0.95), which includes connections of the visual and dorsal attention network (**Figure 2C**). These two PL-states (i.e., clusters c10 and c15 for k=18) are represented in **Figure 2B** as vectors, where each element represents a brain area, and whose value indicates the degree to which the signal in these areas shifts from the main phase direction.

**Figure 2.**
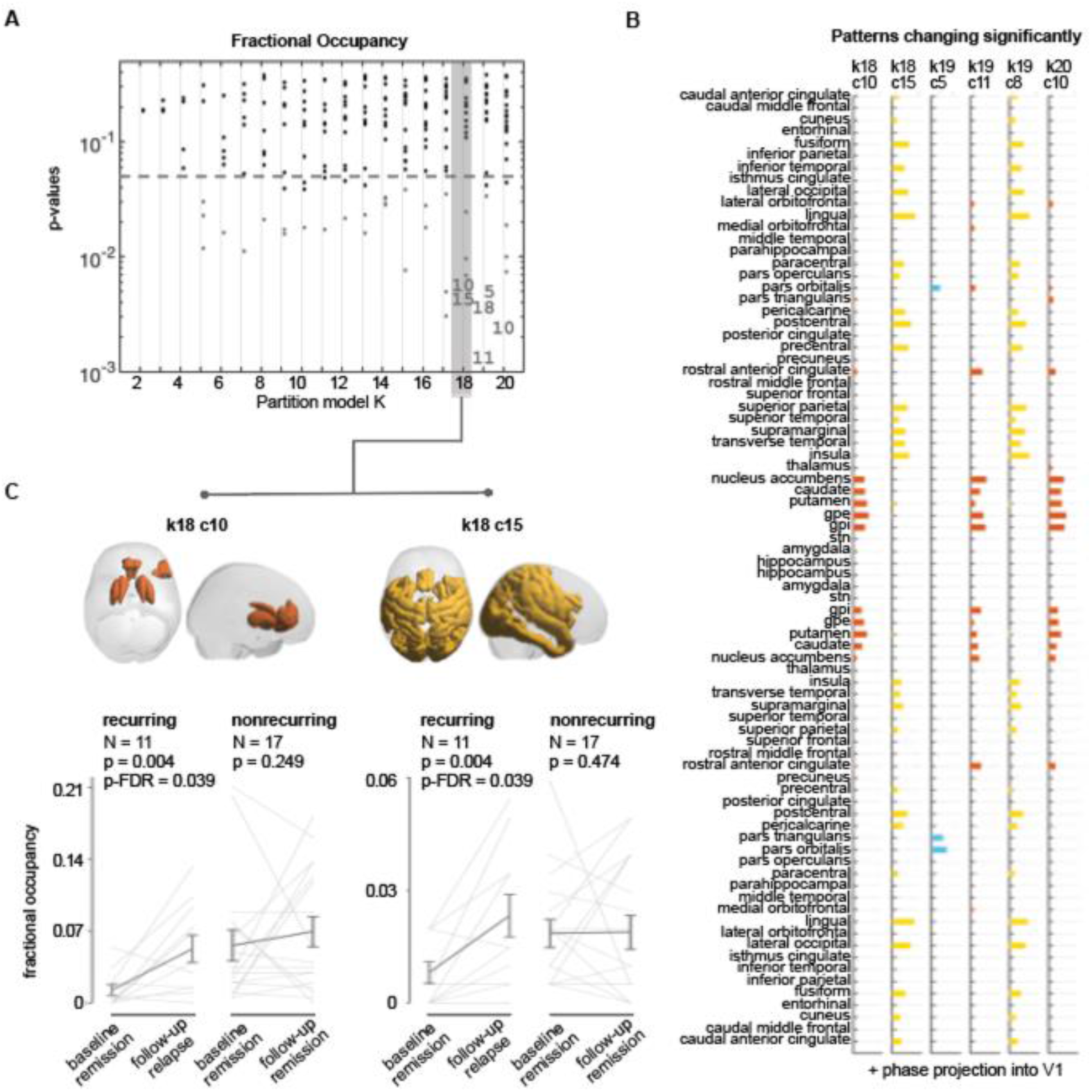
Significant increases in the fractional occupancy of two PL-states during the experience of a new MDD episode. **A)** Statistical significance associated with changes in fractional occupancy from remission to recurrence, in the entire repertoire of PL-states returned by each of the 19 partition models (k2–k20). While most PL-states do not show significant changes (black dots) between baseline (remission) and follow-up (recurrence), for all k>17, two PL-states repeatedly survive FDR corrections (indicated with numbers). PL-states failing to reach the FDR-corrected significance threshold but with p_uncorrected_<0.05 are indicated with gray dots. PL-states are labeled from 1 to k number of clusters considered in each partition model; as a result, variant forms of the same underlying PL-state do not necessarily have the same label in every partition. **B)** Vector representation of the PL-states with a significant (p-FDR<0.05) change in their fractional occupancy from baseline (remission) to follow-up (recurrence). Each bar plot shows the elements in V1, representing fMRI signals of brain regions that become coherent and phase-shifted by more than 90° with respect to the signals in the rest of the brain. States are color-coded according to their similarities; similar forms of these 2 states also appear in partition k19 and k20 (and also in higher partitions see Supplementary Figure S1 for partitions up to k30). **C**) Cortical space representation of the two PL-states, only rendering regions where the phase is shifted more than 90° with respect to the main phase orientation. Underneath each brain plot, graphical representation of the changes in fractional occupancy between baseline and follow-up for the recurring rrMDD patients (i.e. recurrence at follow-up) and the nonrecurring rrMDD patients (i.e., maintaining remission at follow-up). Gray lines represent patient-specific scores, and error-bars represent the mean ± standard error of the mean across subjects. PL = phase-locking.

Significance and effect-sizes were comparable for the k=19 and k=20 clustering solutions. Validating the specificity of our findings in recurring rMDD-patients, we verified that the fractional occupancy of these two states did not change in nonrecurring rMDD-patients. **Supplementary Table S1** shows the fractional occupancy scores and statistics for all PL-states in partition k=18.

### Recurrence-related changes in state lifetime

We also explored within-subject differences regarding state lifetime, that is the duration (in seconds) that a given state occurs. We found two distinct states that significantly changed from baseline to follow-up for several partition models (**Figure 3A**). At lower numbers of PL-state partitions (k=3, 4, 6) a PL-state (k3c3) that consisted of the DMN showed significantly longer lifetimes (shown in gray in **Figure 3B**). Partitioning the data into a high number of clusters (k>8) returned a state characterized by connections within the reward network (shown in brown in **Figure 3B**; k8c4, k10c5, k11c5, k14c7, k17c8, k19c11, k20c10); a variant form of this state was also found to be significantly different from remission to recurrence in terms of fractional occupancy.

**Figure 3.**
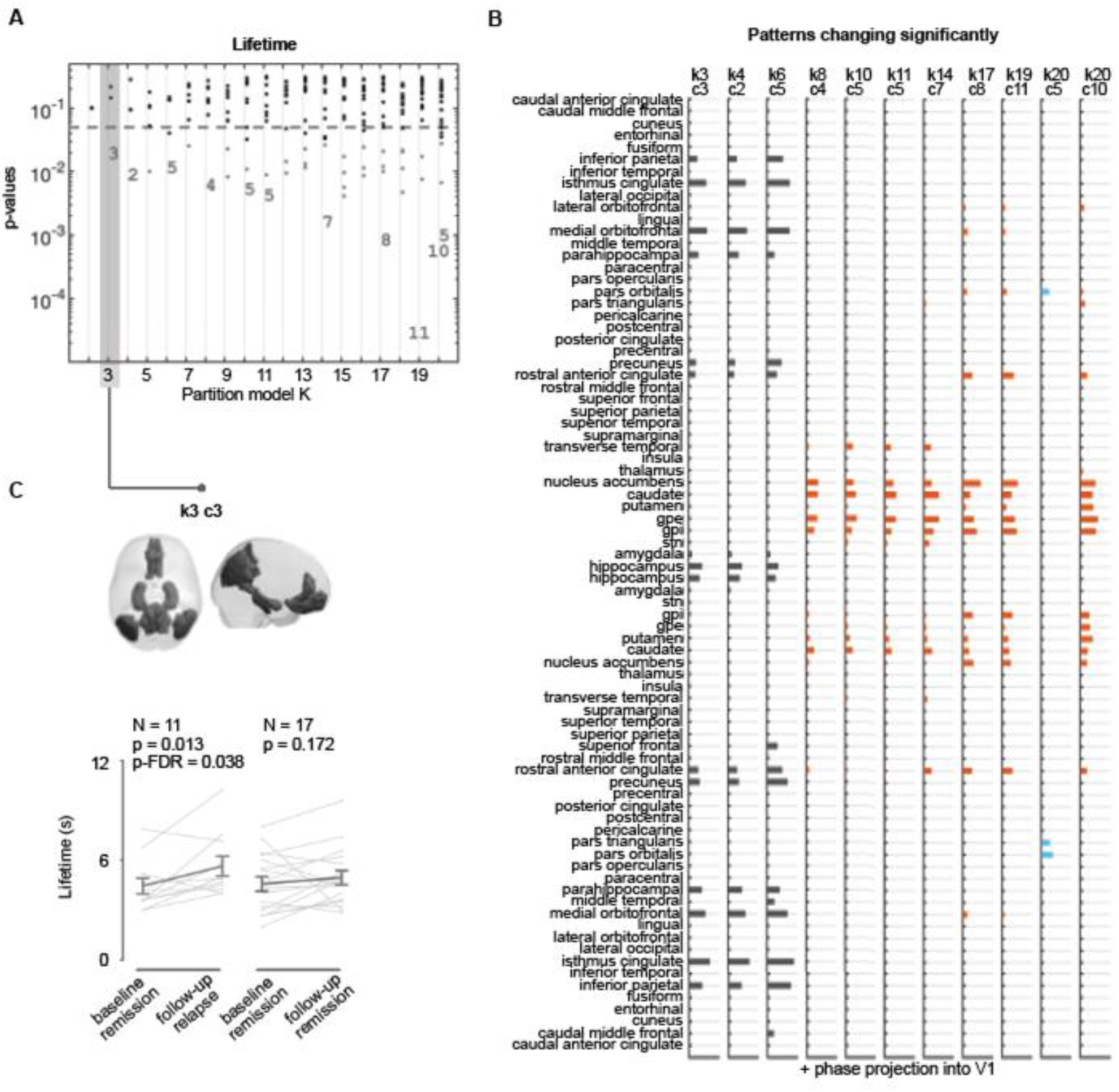
Significant increase in PL-state 3 lifetime during the experience of a new MDD episode. **A)** Statistical significance associated with changes in lifetime from remission to recurrence, in the entire repertoire of PL-states returned by each of the 19 partition models (k2– k20). While most PL-states do not show significant changes between remission and recurrence (black dots), two PL-states repeatedly survive FDR corrections (indicated with numbers). PL-states failing to reach the FDR-corrected significance threshold but with p_uncorrected_<0.05 are indicated with gray dots. PL-states are labeled from 1 to k number of clusters considered in each partition model; as a result, variant forms of the same underlying PL-state do not necessarily have the same label in every partition. **B)** Vector representation of the PL-states with a significant (p-FDR<0.05) change in their lifetime from baseline (remission) to follow-up (recurrence). Each bar plot shows the elements in V1, representing fMRI signals of brain regions that become coherent and phase-shifted by more than 90° with respect to the signals in the rest of the brain. States are color-coded according to their similarities. **C)** Cortical space representation of the centroids of PL-state 3. Only rendering regions phase-shifted more than 90° with respect to the fMRI signals in the rest of the brain. Underneath the brain plot, graphical representation of change in state lifetime between baseline and follow-up for the recurring rrMDD patients (i.e., recurrence at follow-up) and the nonrecurring rrMDD patients (i.e., maintaining remission at follow-up). Gray lines represent patient-specific scores, and error-bars represent the mean ± standard error of the mean across subjects. PL = phase-locking.

The lifetime of a state is calculated as the average number of consecutive frames in that state. The higher the number of clusters considered in a partition model, the shorter the state lifetimes. Given the relatively short length of the recording session (204 volumes) with a TR of 2 seconds, the states resulting from partitions with higher cluster numbers should be interpreted with caution. Therefore, here, we focused on the state returned by the lowest partition: PL-state 3 (k3c3). As **Figure 3C** shows, the mean lifetime of PL-state k3c3 significantly increased from remission to recurrence (*t*(10)=2.44; *p*-FDR=0.038, *d* =0.74). Significance and effect-sizes were comparable for the k=4 and k=6 clustering solutions.

Validating the specificity of our findings in recurring rMDD-patients, we did not find a significant difference in the lifetime of PL-state k3c3 for patients who were not experiencing a recurrent episode at follow-up (*t*(16)=0.96; *p*=0.172). **Supplementary Table S2** shows the lifetime scores and statistics for the states in partition k=3.

### Phase shift of the reward system from the rest of the brain

To further investigate the effects captured with LEiDA, we analyzed how fMRI signals evolve over time in one of the subsystems found to ‘shift in phase’ significantly more often in rMDD-patients in recurrence with respect to baseline. When PL-state k18c10 is on, signals in the regions with a positive sign in this PL-state (corresponding to the basal ganglia and ACC) align together and shift in phase with respect to regions with a negative sign (**Figure 4A**). When comparing the average phase shift between signals in the reward system (basal ganglia, ACC) and the rest of the brain, a significant increase is detected when patients experienced a recurrence of MDD (*t*(10)=2.65, *p*=0.008) (**Figure 4B**). Instead, no significant change was observed for patients who were still in remission at follow-up (*t*(16)=0.14, *p*=0.446). **Supplementary Figure S2** illustrates in more detail the phase shift of the reward system from the rest of the brain.

**Figure 4.**
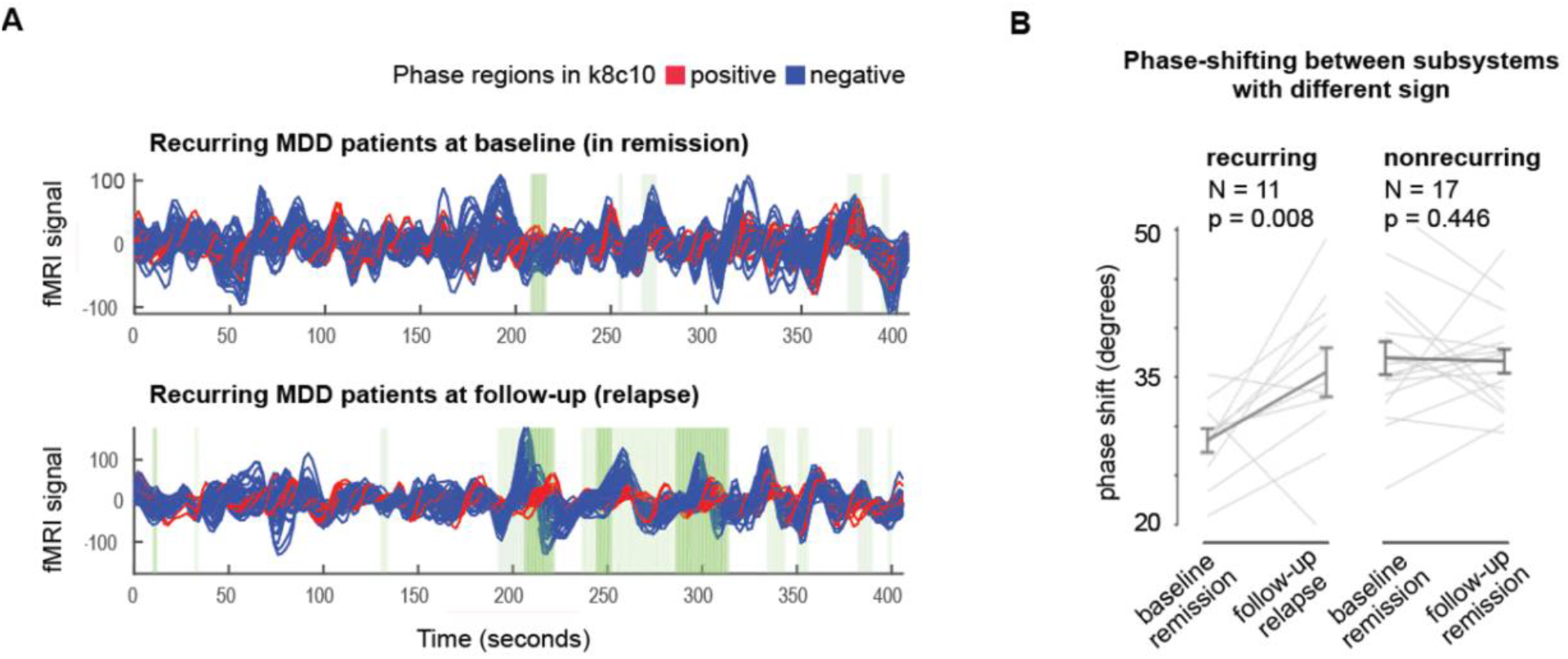
Significant change in phase shift of the reward network from the rest of the brain during the recurrence of an MDD episode. **A)** For illustrative purposes we visualize the fMRI signals in all 80 brain areas during remission (top) and at follow-up, when having a recurrence (bottom) in a single patient. Time-series of the positive community of k18c10 are shown in red, all the remaining brain regions are shown in blue. The 3-dimensional effect is obtained by plotting the Hilbert-transformed ROI signals over time, but the view is shown only from one side. The signals have a real and an imaginary part, which can be perceived as a hidden dimension, on which they evolve while conserving angular momentum. The darker green patches indicate when the state k18c10 was detected by the k-means clustering. The lighter green patches indicate when the leading eigenvector V1(t) is relatively close to the centroid k18c10, to give a notion of when this state may be present, even if not dominant. **B)** Evaluating the average phase-shift between regions of different signs in each PL-state (i.e., the red vs. the blue time series in panel A), we compare this measure across time points and conditions. The recurrent patients (i.e., recurrence at follow-up) are found to significantly increase the phase shift (in degrees) from baseline to follow-up, whereas the nonrecurring rrMDD patients (i.e., maintaining remission at follow-up) did not exhibit any significant change. Gray lines represent patient-specific scores, and error-bars represent the mean ± standard error of the mean across subjects. PL = phase-locking.

### Sensitivity analysis

Since baseline–follow-up differences were tested on the PL-states that were defined solely from the 2 scan sessions of the recurring patients (*n*=11), we tested potential differences between baseline and follow-up on the states derived from the 2 scan sessions of the recurring patients (*n*=17). Indeed, no significant (*p*-FDR<.05) differences were observed for any of the PL-states in any of the 19 partition models (k2–k20) between follow-up (remission) and baseline (maintained remission) for the matched nonrecurring patients (**Supplementary Figure S3**).

## Discussion

In the current study, we investigated dynamic FC changes that occurred in rMDD-patients after transitioning from remission to a recurrence of a depressive episode. We found significant within-subject changes in the fractional occupancy and lifetime duration of three PL-states. Specifically, the reward network (consisting of the basal ganglia and ACC) and a widespread visual-attention network were more likely to occur when suffering from a depressive episode as compared to being in a state of remission. Additionally, the DMN was activated for a longer duration during a recurrence of depression. As a validation, these changes were not seen in a separate sample of matched rMDD-patients scanned twice when remaining in remission with comparable follow-up times, age, sex and IQ. Lastly, we showed post-hoc that the observed changes in the reward network were due to the decoupling of this circuit from the rest of the brain.

Among the neural hallmarks of a major depressive episode are disrupted large-scale brain network communications (Kaiser et al., 2015). Within these networks, dysfunctions of prefrontal-basal ganglia/(para)limbic networks have been linked to a dysfunctional reward system in depressed patients (Martin-Soelch, 2009). In particular, hypo-activity of the striatum has been reported in MDD in reinforcement learning and reward processing task paradigms (Gradin et al., 2011; Hall, Milne, and MacQueen, 2014; Kumar, Waiter, Milders, Reid, and Steele, 2008; Pizzagalli et al., 2009; Robinson, Cools, Carlisi, Sahakian, and Drevets, 2012; Smoski et al., 2009). Also in the overall sample of the DELTA-neuroimaging study, impaired learning signals in the ventral tegmental region differentiated rMDD-patients and controls (Geugies et al., 2019), similar to Kumar et al. (2008).

While these findings suggest a deactivation of the reward system, FC measures report attenuated coupling within the reward network (Li, Friston, Mody, & Hu, 2018; Tol et al., 2013), and relate this to depression severity and MDD symptoms (Satterthwaite et al., 2015; Zhu et al., 2017). In fact, this intrinsic FC impairment appears to be progressive across the course of depression (Liu et al., 2021). Of note, others have shown that the ventral striatum exhibits both hyper- and hypo-connectivity (with the DMN) in MDD (Leaver et al., 2016) and increased information transfer between the OFC and subcortical (limbic) regions in first-episode MDD-patients (Gao, Zou, He, Sun, and Chen, 2016). In line with these latter studies, we found increased fractional occupancy of a state involving the reward network whilst experiencing a depressive episode.

Note that the increased fractional occupancy of this state may not directly translate to increased activity or connectivity of the reward network. By introducing a new “decoupling metric”, we further demonstrated that this state was characterized by a consistent de-phasing of the reward network from the rest of the brain, suggesting network decoupling and segregation during recurrence. Similarly, other studies have pointed to a desynchronization in striatal-frontal connectivity (Leaver et al., 2016), synergistic functional decoupling in the reward network correlated with anhedonia (Gong et al., 2018), and increased FC fluctuations within thalamic and basal-ganglia networks (Long et al., 2020).

A desynchronization or decoupling of neural networks may likely be an indicator of the functional pathophysiology of MDD, extending to other networks such as the DMN and the visual-attention network. In fact Marchetti et al. (2012) initially proposed an imbalance between task-positive and task-negative elements of the DMN as a risk factor for depression recurrence. Later studies in MDD have pointed to altered neural networks and connections with and within the DMN (Li et al., 2020; Liu et al., 2017), as well as excessive fluctuations in FC in DMN (Long et al., 2020). Others have shown a persistent reduction of low-frequency fluctuations (representing intrinsic local neuronal activities) in the precuneus and posterior cingulate cortex, despite treatment or achieved remission (Wang et al., 2020) - regions considered the functional core of the DMN (Utevsky, Smith, and Huettel, 2014). Such a MDD-related disruption in network communication in the DMN has also recently been reported with respect to rich-club organization (the basis for long-range, high-capacity signaling) (Liu et al., 2020). Dominance of DMN activity has been associated with MDD recurrence and related to rumination (Hamilton et al., 2011; Lythe et al., 2015). Similarly, in this study we found increased recruitment (lifetime) of the DMN during a depressive state versus previous remission, reflecting perhaps the incremental nature of DMN dominance and a particular failure to control internally-focused thoughts in recurrence.

Other studies also point to disrupted communication in or with visual-attention circuits. For example, stronger connectivity of attention regulation circuits to DMN has been implied to relate to better responsivity to internally generated self-relevant thoughts rather than outside stimuli (Knyazev et al., 2018). Stability of FC (vs. more frequent and rapid shifts) in visual-attention regions, such as the ACC, calcarine sulcus, and middle occipital gyrus has been shown to predict improvement in depressive symptoms in MDD (Li et al., 2021). Here, we also found that the visual-attention circuit was more likely to occur during a recurrence, compared to remission, which could represent attempts to regulate and overcome depressive phenomena in patients experiencing a recurrence.

Importantly, what we show with the results of the current study, is that network reconfiguration takes place within individuals when they transition from a state of remission to a depressive episode. As such, implementing dynamic connectivity approaches in clinical populations could give insight into changes that occur across time in individuals, especially during vulnerable periods (e.g., recurrence of a depressive episode). Future work should investigate how these neural measures could be used to improve recurrence prevention and what would be the optimal time to administer treatment. The finding of increased segregation of certain brain networks during a depressive episode might help to identify ways of improving flexibility and integration of the implicated networks. For example, electroconvulsive therapy has been shown to impact ventral striatum connectivity – a region deemed to be a key structure contributing to network desynchronization in MDD (Leaver et al., 2016). Identifying the source of the transition from integrated to more segregated neural states could, therefore, be used in the future as a target for tailoring interventions based on individualized risk assessments.

### Interpretational Issues

Several limitations should be considered with respect to the results of this study. Arguably, the reliability of the presented inferences is limited by the low sample size (n=28). However, this should be weighed against the uniqueness and specificity of the sample – antidepressant-free patients with at least two past episodes of MDD, scanned during remission, followed-up and then scanned again while in a depressive episode, as well as a matched comparison group of MDD participants without a recurrence when scanned again. While these findings should be taken tentatively and replicated in future longitudinal studies, they provide an initial insight into the neural biomarkers changing in the perspective of recurrence.

Although the nonrecurring patients were in remission during the follow-up scan, a possibility remains that they had a depressive episode at a later time. The fact that we find differences in PL-state profiles in the recurring, but not in the nonrecurring group, are associated with the present experience of acute depressive symptoms. A full factorial comparison of recurrent and non-recurrent patients at baseline (and changes over time) was not possible given the group sample sizes. Future investigations may identify how specific symptomatology, such as anhedonia (Servaas et al., 2017) relates to changes in these FC dynamics.

It could be argued that the networks we identified could be restricted due to the number of states (k) that best represents FC dynamics. Here, we assessed our results for a wide range of k (from 2 to 20), and further demonstrated the consistency and significance of our findings across several choices of k. Although it can be argued that the networks were constrained by the parcellation atlas that was used, in contrast to (Figueroa et al., 2019) who used the Anatomical Automated Labeling atlas, we used a clinically relevant, custom-made ‘DBS80’ parcellation, designed specifically for DBS studies.

Although the new “decoupling metric” was implemented based on results obtained from LEiDA, these two approaches provide novel insights regarding underlying differences in fractional occupancy of the reward network. As such, the decoupling metric is not only a confirmation, but also an extension of the results obtained using LEiDA. With LEiDA, only the epochs when a particular pattern is dominant (‘represented by the leading’ eigenvector) are taken into account, whereas decoupling is computed over the entire time series. This approach relies on the hypothesis that phase relationships between areas are critical. For example, one area consistently activates a bit later than the other – as such these areas would be coupled, but not in-phase, as typically captured by co-activation measures. Using this new metric, we can quantify the mean phase shift (or angular difference) between baseline and follow-up and, therefore, investigate whether depression recurrence is associated with a consistent de-phasing of one network from the rest of the brain.

## Conclusion

With a unique repeated measures design of remitted depressed patients followed-up for 2.5 years while scanned in remission and in a recurrence episode, this study provides first evidence of alterations in the brain’s dynamical repertoire that are specifically associated with the transition from remission to recurrence of a depressive episode. These results highlight the role of widespread reconfiguration of networks known to be implicated in MDD, such as the DMN, reward and attention circuits. Crucially, we show that these changes occur within individuals and as such provide a promising avenue for emerging personalized treatment.

## Data Availability

Data is available upon request via the Donders Repository (https://data.donders.ru.nl/). Data can be provided by the Donders Institute for Brain, Cognition, and Behaviour pending scientific review and a completed data transfer agreement in collaboration with AmsterdamUMC. Requests for the data should be submitted to HR. The MATLAB scripts used to analyse the data in this study are publicly available at https://github.com/sonsolesalonsomartinez/rrMDD.

