## Supplementary Material for "Segregation of dynamic resting-state reward, default mode and attentional networks after remitted patients transition into a recurrent depressive episode"

**Supplementary Table S1.** Fractional occupancy scores of states in partition k18 and associated statistics

| | Recurring rrMDD-patients (i.e., relapse at FU; N=11) | | | | Nonrecurring rrMDD-patients (i.e., no relapse at FU; N=17) | | |
|---|---|---|---|---|---|---|---|
| state | Baseline mean ± std | Follow-up mean ± std | p | pFDR | Baseline mean ± std | Follow-up mean ± std | p |
| c1 | 0.35 ± 0.25 | 0.31 ± 0.22 | 0.282 | 0.391 | 0.36 ± 0.18 | 0.36 ± 0.14 | 0.485 |
| c2 | 0.14 ± 0.09 | 0.13 ± 0.10 | 0.458 | 0.459 | 0.19 ± 0.09 | 0.16 ± 0.07 | 0.133 |
| c3 | 0.11 ± 0.08 | 0.08 ± 0.05 | 0.142 | 0.391 | 0.07 ± 0.05 | 0.07 ± 0.06 | 0.324 |
| c4 | 0.07 ± 0.06 | 0.03 ± 0.04 | 0.012 | 0.056 | 0.04 ± 0.02 | 0.04 ± 0.04 | 0.400 |
| c5 | 0.06 ± 0.07 | 0.04 ± 0.03 | 0.197 | 0.391 | 0.03 ± 0.03 | 0.03 ± 0.03 | 0.411 |
| c6 | 0.04 ± 0.04 | 0.04 ± 0.04 | 0.312 | 0.401 | 0.05 ± 0.04 | 0.03 ± 0.03 | 0.082 |
| c7 | 0.03 ± 0.06 | 0.05 ± 0.12 | 0.231 | 0.391 | 0.01 ± 0.01 | 0.01 ± 0.02 | 0.193 |
| c8 | 0.03 ± 0.03 | 0.05 ± 0.05 | 0.267 | 0.391 | 0.03 ± 0.04 | 0.05 ± 0.04 | 0.040 |
| c9 | 0.03 ± 0.03 | 0.03 ± 0.04 | 0.459 | 0.459 | 0.03 ± 0.02 | 0.02 ± 0.01 | 0.045 |
| c10 | 0.01 ± 0.02 | 0.05 ± 0.04 | 0.004 | **0.039** | 0.06 ± 0.06 | 0.07 ± 0.06 | 0.249 |
| c11 | 0.03 ± 0.03 | 0.03 ± 0.03 | 0.431 | 0.459 | 0.03 ± 0.04 | 0.02 ± 0.03 | 0.205 |
| c12 | 0.01 ± 0.01 | 0.05 ± 0.04 | 0.009 | 0.053 | 0.03 ± 0.02 | 0.04 ± 0.04 | 0.038 |
| c13 | 0.01 ± 0.01 | 0.03 ± 0.04 | 0.155 | 0.391 | 0.03 ± 0.03 | 0.02 ± 0.02 | 0.176 |
| c14 | 0.01 ± 0.02 | 0.02 ± 0.04 | 0.175 | 0.391 | 0.01 ± 0.02 | 0.02 ± 0.02 | 0.202 |
| c15 | 0.01 ± 0.01 | 0.02 ± 0.02 | 0.004 | **0.039** | 0.02 ± 0.02 | 0.02 ± 0.02 | 0.474 |
| c16 | 0.02 ± 0.03 | 0.01 ± 0.01 | 0.032 | 0.115 | 0.01 ± 0.02 | 0.01 ± 0.01 | 0.342 |
| c17 | 0.02 ± 0.02 | 0.01 ± 0.01 | 0.263 | 0.391 | 0.01 ± 0.01 | 0.01 ± 0.02 | 0.370 |
| c18 | 0.01 ± 0.01 | 0.02 ± 0.05 | 0.456 | 0.459 | 0.00 ± 0.01 | 0.01 ± 0.01 | 0.139 |

**Supplementary Table S2.** Lifetime scores (in seconds) of states in partition k3 and associated statistics

| | **Recurring rrMDD-patients (i.e., relapse at FU N=11)** | | | | **Nonrecurring rrMDD-patients (i.e., no relapse at FU; N=17)** | | |
|---|---|---|---|---|---|---|---|
| state | Baseline mean ± std | Follow-up mean ± std | p | pFDR | Baseline mean ± std | Follow-up mean ± std | p |
| c1 | 28.9 ± 22.1 | 33.6 ± 43.5 | 0.34 | 0.34 | 33.7 ± 28.5 | 27.0 ± 13.3 | 0.15 |
| c2 | 5.7 ± 2.3 | 4.9 ± 2.3 | 0.23 | 0.34 | 4.8 ± 1.7 | 4.5 ± 2.0 | 0.32 |
| c3 | 4.4 ± 1.6 | 5.6 ± 1.9 | 0.01 | **0.04** | 4.6 ± 1.8 | 4.9 ± 1.7 | 0.17 |

**Supplementary Figure S1**

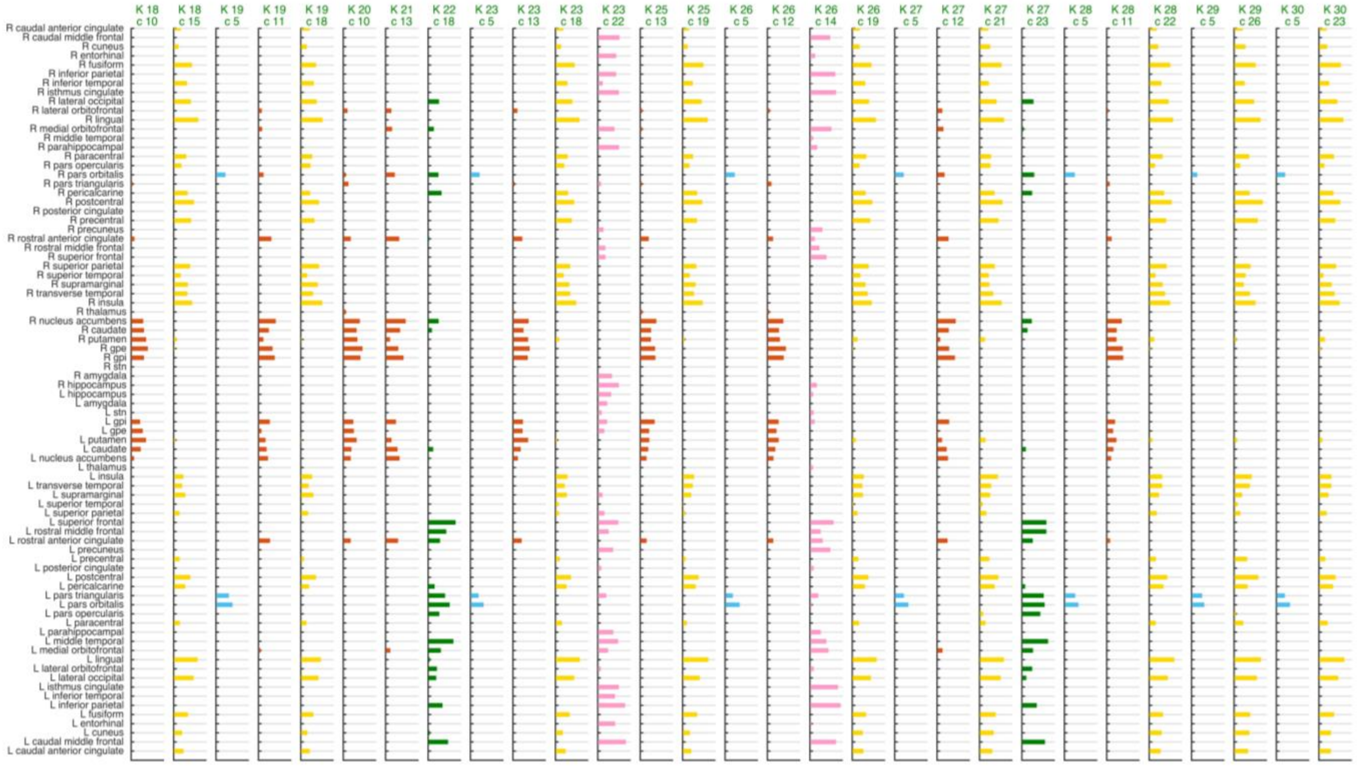


Vector representation of the cluster centroids of PL-states with a significant (p-FDR<0.05) change in their fractional occupancy from baseline (remission) to follow-up (recurrence) when the clustering solution is expanded to 30 clusters. FDR-corrected significance is based on permutation paired t-test (*N*=11). Each bar plot shows the elements in $V_1$ representing BOLD signals of brain regions that become coherent and phase-shifted by more than 90° with respect to the BOLD signals in the rest of the brain. *K* indicates a partition solution into *k* clusters; *c* indicates the numbering of the PL-state. Note that for each partition model (*k*), the PL-states (*c*) are labelled from 1 to *k* number of clusters considered in each partition model; as a result, variant forms of the same underlying PL-state do not necessarily have the same label in every partition. Therefore, bar plots of the same color represent similar forms of the underlying PL-state, showing stability of the PL-state across different clustering solutions.

PL = phase-locking.

**Supplementary Figure S2**.

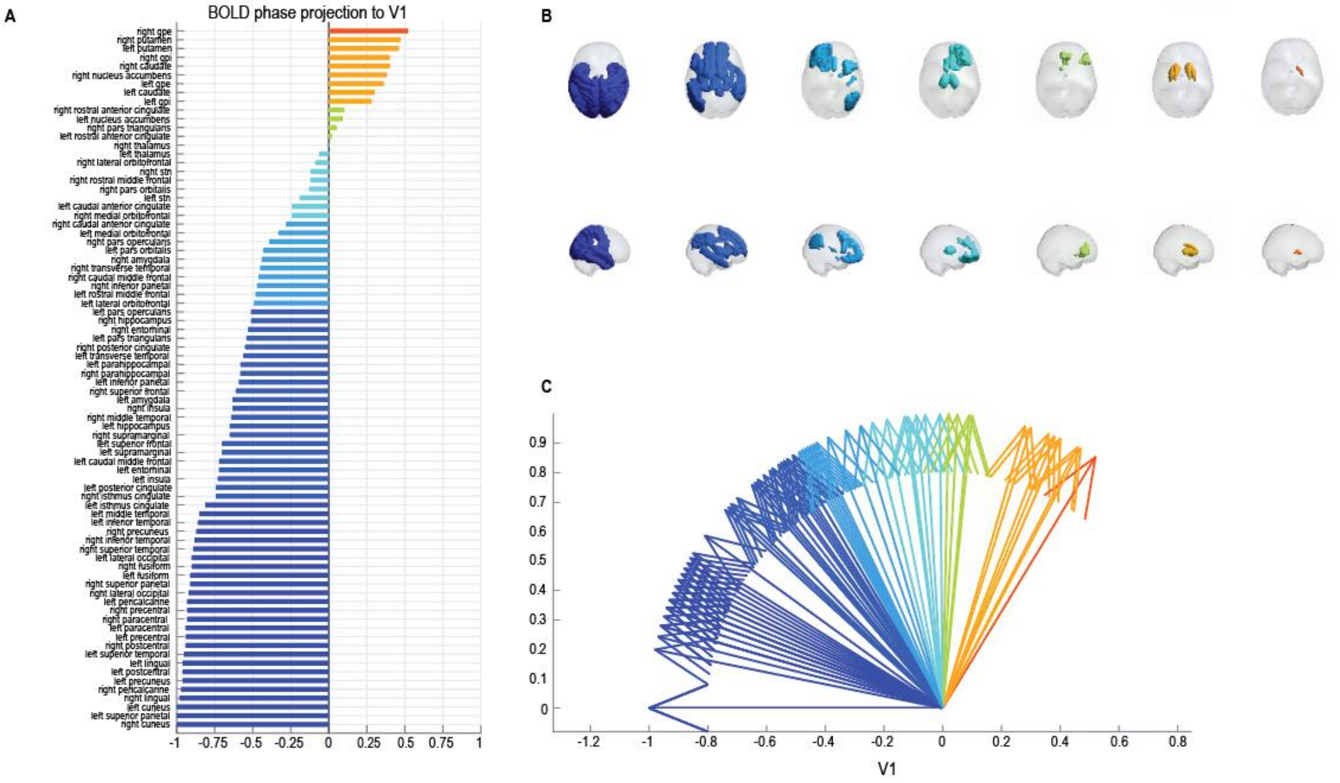


Brain activity during MDD episodes is characterized by a decoupling of the reward system from the rest of the brain. **A** Bar plot shows the elements of the leading eigenvector *V1* of PL-state 10 (k18c10) ordered by strength. Regions of the reward system are depicted in red, orange and green; the BOLD signals of these regions become phase-shifted by more than 90° with respect to the BOLD signals in the rest of the brain (cyan-blue regions). **B** Regions of the same color are rendered in a brain surface. **C** The direction of the arrows indicates the sign of the corresponding element of the Leading eigenvector $V_1$.

**Supplementary Figure S3.**

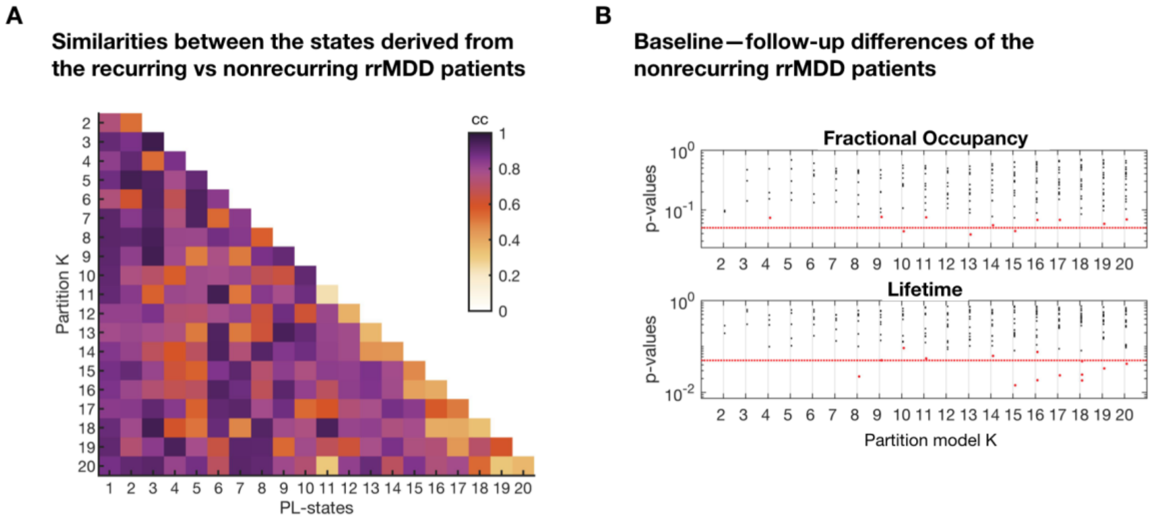


**A** Similarity matrix shows the correlation coefficients between the PL-states that resulted from clustering the data from the recurring rrMDD patients and the PL-states that resulted from clustering the data from the nonrecurring rrMDD patients. **B** Statistical significance associated with changes in fractional occupancy (top) and lifetime (bottom) between baseline (remission), and follow-up (maintained remission) of the nonrecurring rrMDD patients. Note that contrary to the figures reported in the main text, here, differences between baseline and follow-up scans of the nonrecurring patients were tested on the PL-states defined directly on the data from the nonrecurring patients. Most PL-states do not show significant changes between remission and recurrence (black dots), no PL-states survive FDR corrections (green). PL-states failing to reach the FDR-corrected significance threshold but with $p_{uncorrected}$<0.05 (red dotted line) are indicated in red.
